# Understanding the Collective Responses of Populations to the COVID-19 Pandemic in Mainland China

**DOI:** 10.1101/2020.04.20.20068676

**Authors:** Haoyi Xiong, Ji Liu, Jizhou Huang, Siyu Huang, Haozhe An, Qi Kang, Ying Li, Dejing Dou, Haifeng Wang

**Affiliations:** Baidu Inc., Beijing, 100085, China

## Abstract

Timely information acquisition and stay-at-home measures have been considered as two effective steps that every person could take to help contain the coronavirus (COVID-19) pandemic. From the perspectives of information and mobility, this work aims at evaluating to what degree the massive population has responded to the emergencies of the COVID-19 pandemic in China. Using the real-time and historical data collected from the Baidu Maps and Baidu search engines, we confirm the strong correlation between the local pandemic situation in every major Chinese city and the population inflows from Wuhan between 1 January and 23 January 2020. We further evidence that, in cities under more critical situations, people are likely to engage COVID-19-related searches more frequently, while they are not likely to escape from the cities. Finally, the correlation analysis using search and mobility data shows that well-informed individuals are likely to travel less, even while the overall travel demands are low compared to the historical records. Partial correlation analysis has been conducted to test the significance of these observations with respect to other controlling factors.

## Introduction

The coronavirus disease (COVID-19) has become a global emergency that is currently rampaging throughout the whole world^1,2^. Wuhan (Hubei Province, China) was the first Chinese major city which had a COVID-19 outbreak. To deal with the critical situation of the COVID-19 outbreak in China, a series of quarantines, isolation, and travel restrictions in municipal areas, provincial areas, and nationwide have been placed by Chinese authorities, while the outcomes of these interventions have been examined in^3–5^. On the other hand, there are a great number of studies^6–13^ that have demonstrated feasibility to leverage mobile applications and search engines, such as Baidu Migration^1^ and Baidu COVID-19 Page^2^, to acquire the real-time information on pandemics, including trends of human mobility, confirmed cases, and death tolls for Chinese cities.

In this work, we aim at evaluating to what degree mobility controls and information acquisitions would affect spread and infection of COVID-19 in every major Chinese city. Further, we wish to understand in which way the three factors — mobility controls, information provisions/acquisitions, and infections — would interplay with each other. More specifically, we would like to understand following issues using realistic data.

- *To what degree did the population inflows from Wuhan (before quarantine) affect the pandemic outbreaks of COVID-19 in each city of China?* This research issue has been studied^12,13^ using the migration scale index released by Baidu Migration Open Data, where the scale index has been normalized using the past records due to the privacy concerns. We propose to provide solid results using exact figures for every major Chinese city.
- *Would people living under more critical infection situations conduct more pandemic-related internet searches?* We hypothesized that people might need to acquire information more frequently (as was seen in the collective responses to the emergencies^14^ and panics^7^) when their pandemic situations become worse. We therefore propose to study the correlation between the per capita pandemic search frequency and the number of local infection cases and local infection rate (the average number of confirmed infection cases per 10 millions) for every major Chinese city.
- *Would people escape from the cities that have more infection cases or higher infection rates?* Again, considering the collective mobility patterns of populations evidenced in crisis^15,16^, we thus hypothesized that people might want to leave the cities with more critical infection situations. We therefore propose to study the correlation between the outflows from every major Chinese city and the number of local infection cases and local infection rate.
- *Would people more frequently searching for pandemic-related information travel less?* With more pandemic-related information acquired through searches, individuals turn out to be better informed. We hypothesized that well-informed individuals would travel less to lower the risk of infection. We would like to understand whether the people with higher per capita search would reduce their travels.

Compared to the existing research^6,12,13^, we particularly focused on the scale of city as a potential controlling variable that would affect the results of correlation analysis. It is reasonable to assume that a larger city with a greater population, rather than the smaller ones, might incorporate more travels, more infections, and even more per capita internet search activities. Furthermore, we are the first to study COVID-19-related internet search behavior subject to the pandemic and mobility issues in Mainland China.

## Results

We collected four datasets (please see also the Methods section), which characterized the mobility and search behaviors related to COVID-19 from more than 1 billion users of Baidu search and Baidu Maps in Mainland China. We then carried out the data-driven analysis using the four dataset to obtain four unique observations (summarized in Table 1) as follows.

**Table 1.** Overall Results of Correlation Analysis

| Correlations | Controlling | Coeff. (R) | p-value |
| --- | --- | --- | --- |
| *Result I* |  |  |  |
| #Confirmed cases vs. #Inflows from Wuhan | N/A | 79.58% | < 0.0001 |
|  | #Populations | 73.37% | < 0.0001 |
| Infection rates vs. #Inflows from Wuhan | N/A | 33.68% | < 0.0001 |
|  | #Populations | 38.92% | < 0.0001 |
| *Result II* |  |  |  |
| Per capita COVID-19 search vs. #Confirmed cases | N/A | 29.40% | < 0.0001 |
|  | #Populations | 13.46% | < 0.05 |
| Per capita COVID-19 search vs. infection rates | N/A | 42.03% | < 0.0001 |
|  | #Populations | 41.28% | < 0.0001 |
| *Result III* |  |  |  |
| Outflow recovery rates vs. #Confirmed cases | N/A | -22.16% | < 0.0001 |
| Outflow recovery rates vs. Infection rates | N/A | -17.42% | < 0.01 |
| *Result IV* |  |  |  |
| Outflow recovery rates vs. Per capita COVID-19 search | N/A | -30.96% | < 0.0001 |
|  | #Populations | -28.58% | < 0.0001 |
|  | #Confirmed cases | -26.23% | < 0.0001 |
|  | Infection rates | -26.47% | < 0.0001 |

### Significant correlations have been evidenced between the population inflows from Wuhan and local infections in major Chinese cities

*We have evidenced the significance of the correlations between the population inflows from Wuhan and local infections (both the number of confirmed cases and the infection rate) for major Chinese cities in the study*. The Pearson correlation between the number of confirmed cases of every city and the number of Baidu Maps users migrated to the city from Wuhan (during 1 to 23 Jan 2020) is *R*^***^ *=* 79.58% (*N* = 307 and *p*-value= 2.03 × 10^−68^ < 0.0001). Similar correlation results have been found in^12,13^. However, we considered that the scale of the city would be a threat to the validity of this observation, as a larger city would attract more population inflows, while a larger city with the greater population would experience more infections.

We therefore tested the significance of correlations between the local infection rate (number of confirmed cases/population of the city) and the population inflows from Wuhan (during 1 to 23 Jan 2020), where we evidenced the significance in the correlations as *R*^***^ *=* 33.68% (*N* = 307 and *p*-value=1.41 × 10^−9^ < 0.0001). Furthermore, we carried out partial correlation analysis between the number of confirmed cases and the population inflows from Wuhan with the effects of the city population size (a controlling variable) removed. The partial correlation analysis gave a strong correlation with significance, such that *R*^***^ *=* 73.73% (*N* = 307 and *p*-value= 3.96 × 10^−53^ < 0.0001). We thus can conclude that no matter whether the city size is large or small, the number of confirmed cases has a significant positive correlation with the population inflows from Wuhan (during 1 to 23 Jan 2020). Please see also in Figure 1 for the volumes of population inflows from Wuhan evolving over time and Figure 2 for the correlations in details.

**Figure 1.**
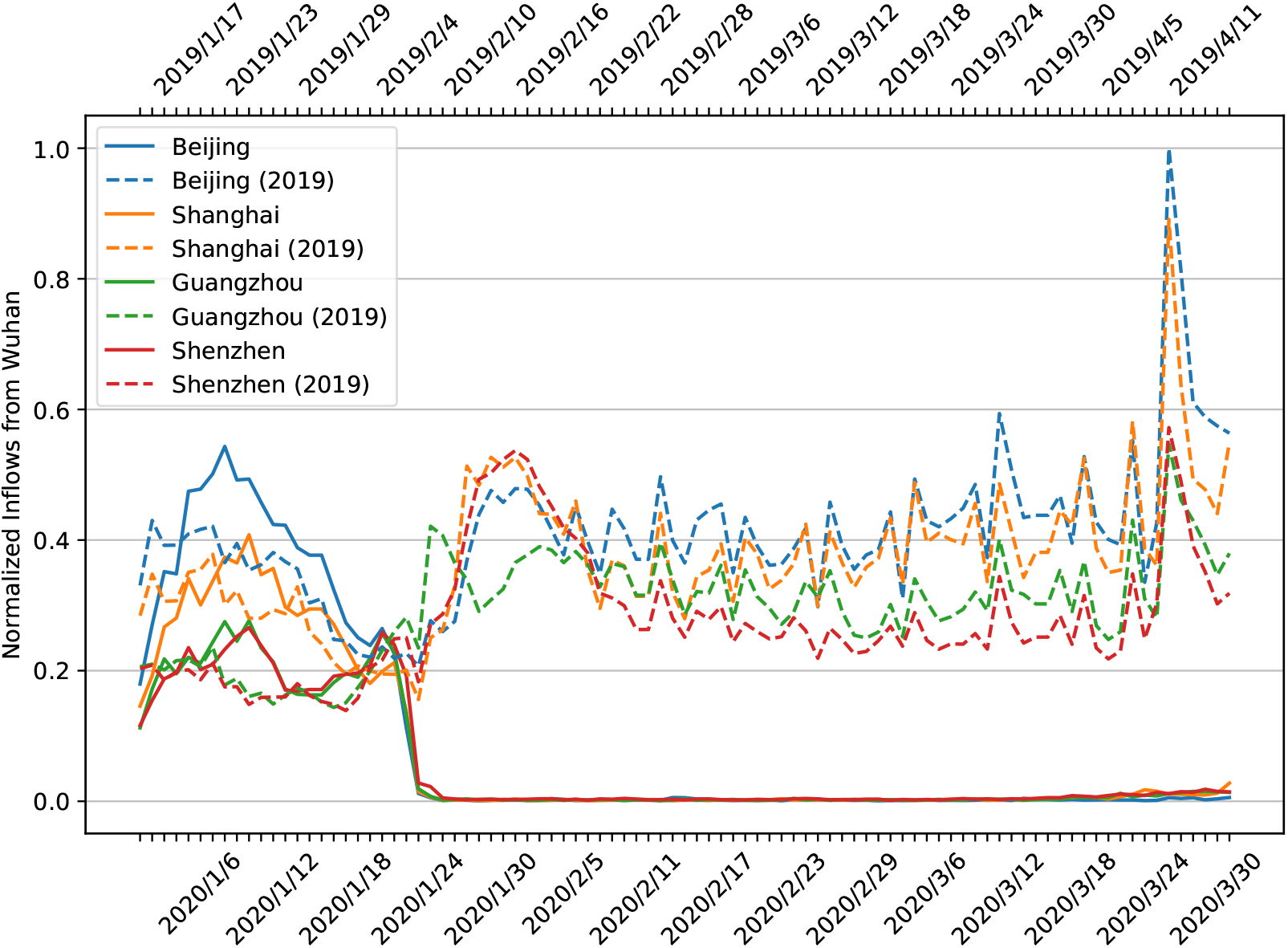
The comparisons of normalized volumes of population inflows from Wuhan between 2020 and 2019. We compared the normalized daily volumes of population inflows from Wuhan to Beijing, Shanghai, Guanghzou, and Shenzhen from 1 Jan 2020 to 31 Mar 2020 with the population inflows in the comparable time period around the Chinese New Year of 2019. A significant reduction could be observed in the volumes of 2020 caused by the quarantine of Wuhan since 23 Jan 2020. The plots have been normalized proportionally using the maximum volume of population inflows (from Wuhan to Beijing in 7 Apr 2019, Sunday and the end of Qingming Festival). Please see also the Supplementary Materials for details about the peak and data normalization.

**Figure 2.**
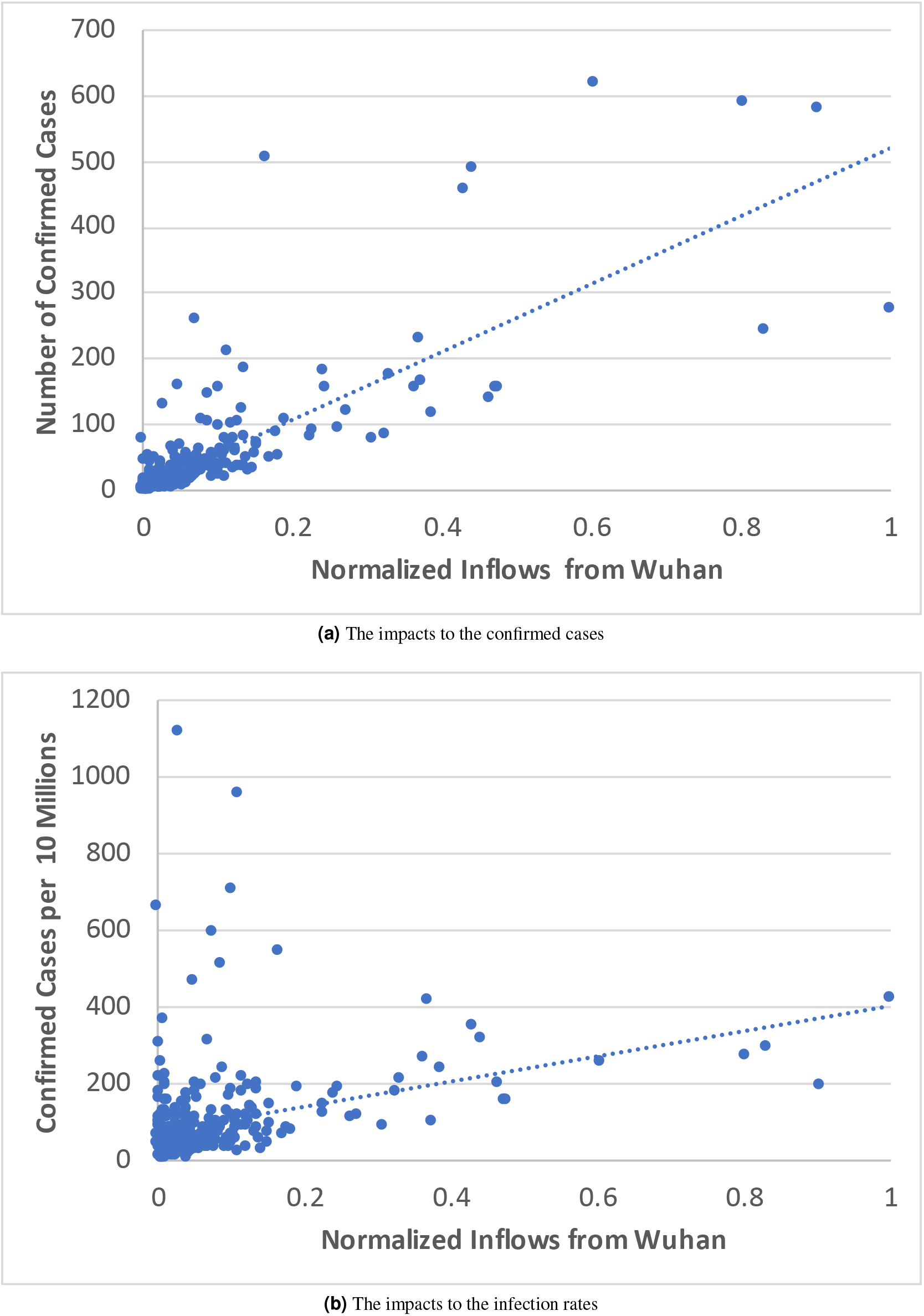
Understanding the impact of population inflows from Wuhan (during 1 to 23 Jan 2020) to the local pandemic situation (by 31 Mar 2020) in every major Chinese city. (Significant positive correlations have been found between both the number of confirmed cases and the local infection rate of every city and the population inflows from Wuhan.)

### Significant correlations have been evidenced between the per capita COVID-19-related search activities and local infections in major Chinese cities

*We have evidenced the significance of the correlations between the per capita COVID-19-related search frequency and local infections (both the number of confirmed cases and the infection rate) for Chinese cities in the study*. We performed correlation and partial correlation analysis using the COVID-19-related search data and the infection data. We found a significant positive correlation between the per capita COVID-19-related search frequency and the number of confirmed cases in every studied city, such that *R*^***^ = 29.40% with *N =* 307 and *p*-value= 1.55 × 10^−7^ < 0.0001. Similarly, with respect to the size of city, we found the correlations between the local infection rate and the per capita COVID-19-related search frequency are even stronger with *R*^***^ = 42.03% (*N* = 307 and *p*-value= 1.42 × 10^−14^ < 0.0001). The partial correlations with the city population size as the controlling variable are significant as well, such that *R*^*^ *=* 13.46% (*N* = 307 and *p*-value= 0.0171 < 0.05) with the number of confirmed cases and *R*^***^ *=* 41.28% (*N* = 307 and *p*-value= 4.65 × 10^−14^ < 0.0001) with the local infection rates.

We thus can conclude that for every city in the study the per capita COVID-19-related search frequency has significant positive correlation to the number of confirmed cases, as well as to the local infection rates. We believe it is due to the natural response to the fear and massive panics^7^. The overall trends of COVID-19-related searches have stepped down since 1 Feb 2020. It might reflect a decrease in the magnitude of public panic. Please see also Figure 3 for the volumes of COVID-19-related searches evolving over time, and Figure 4 for the visualization of the correlations.

**Figure 3.**
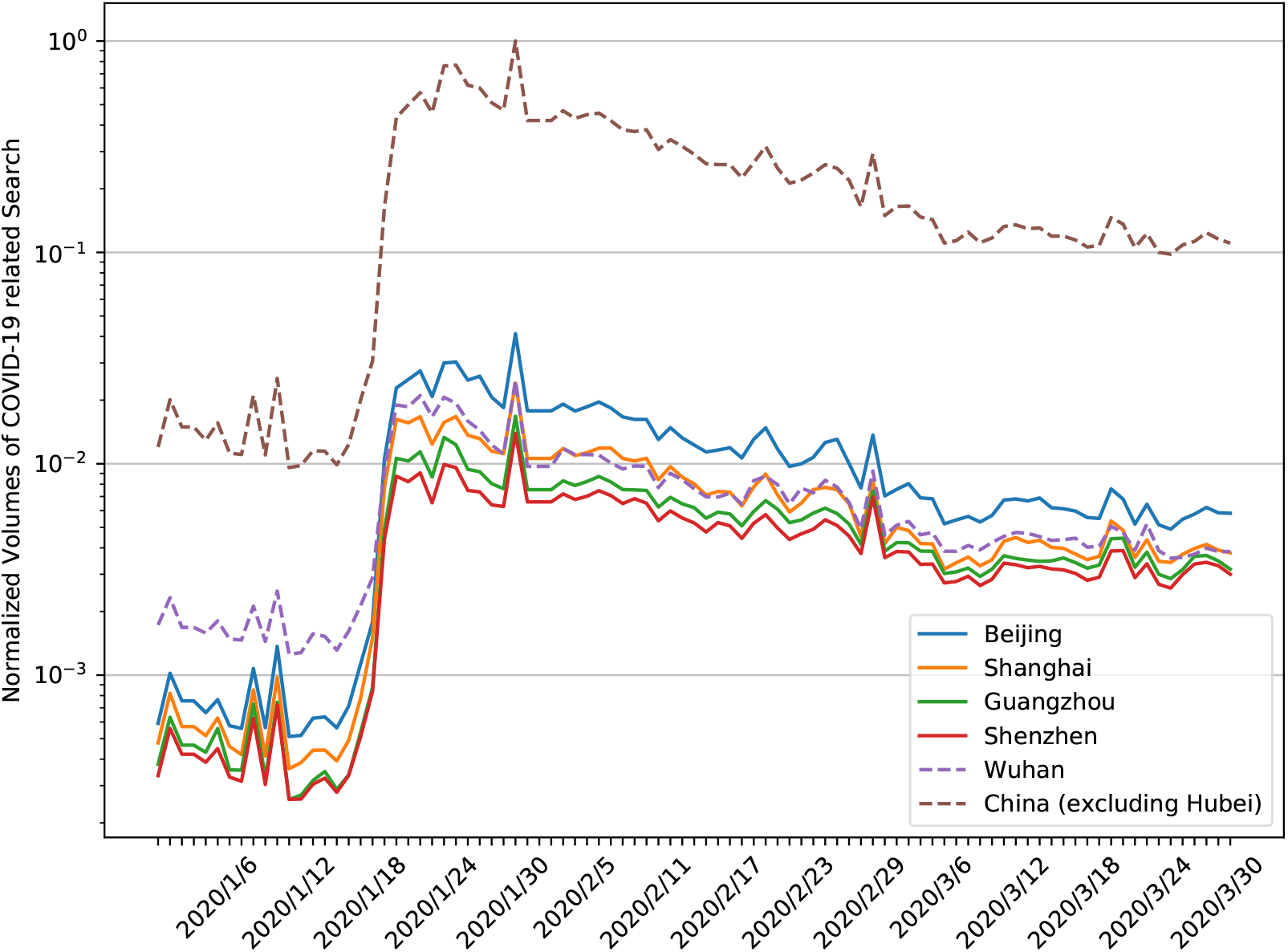
The normalized volumes of COVID-19-related searches over time (in logarithmic scale). We normalized the plots proportionally using the maximum volume of the search (the national peak arrived on 31 Jan 2020 — the date that the first patient cured of novel coronavirus in Wuhan was discharged from the hospital). Wuhan, which has a smaller population than the other four cities, contributed even higher volumes of related searches in the early of January 2020. It might reflect the local infections in the early stage, as well as the collective responses of populations. Nationwide, compared to the peak time, the overall volumes of COVID-19-related searches have dropped to 10% by the end of March 2020. It is possibly due to the collective responses of populations to the relative local containment of the pandemic.

**Figure 4.**
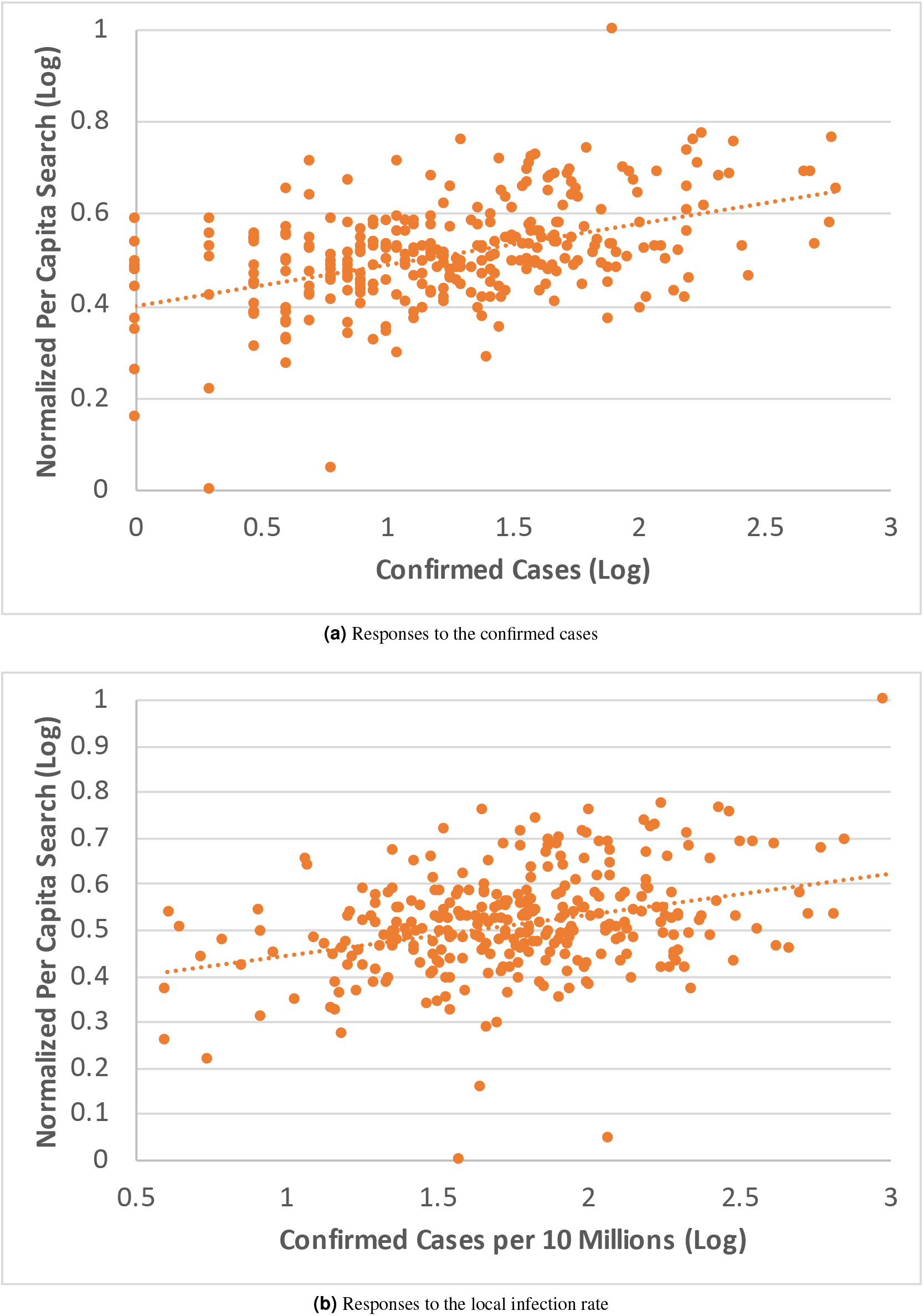
Understanding the COVID-19-related search behaviors as the collective responses of populations to the infectious situation in every city. (Significant positive correlations have been found between the per capita COVID-19-related search frequency and both the number of confirmed cases and the local infection rate of every city.)

### Negative correlations have been evidenced between the population outflows and local infections for major Chinese cities

*We have evidenced the significance of the negative correlation between the outflow populations and local infections (both the number of confirmed cases and the infection rate) for major Chinese cities in the study*. To measure the outflow traffic rates of different cities with various scales, we proposed **outflow recovery rates** that *normalize the outflow traffic rates of every studied Chinese city from 23 Jan to 31 Mar 2020 using the comparable traffic rates in the same period of 2019 Chinese New Year as our basis*. Among all 307 cities in the correlation study, we found the outflow recovery rates range between 17.9% to 66.8% while more than 90% of cities had outflow recovery rates lower than 50%.

We hypothesized that people would try harder to escape the cities with more infections. Therefore, for every city in the study, we correlated the outflow recovery rate with the number of confirmed cases, as well as the outflow recovery rate with the local infection rate, where we obtained Pearson correlation coefficients of *R*^***^ *=* −22.16% (*N =* 307 and *p*-value= 9.00 × 10^−5^ < 0.0001) and *R*^**^ *=* −17.42% (*N* = 307 and *p*-value= 0.002 < 0.01). The correlation analysis result suggests that people under the more critical situations are more likely to refrain from travelling out (fewer travels under higher infections), as the correlation is significantly negative between the outflow recovery rates and infections. Please see also in Figure 5 for the visualization of the correlations.

### Negative correlations have been evidenced between the population outflows and the per capita COVID-19-related search activities

*We have evidenced the significance of the negative correlation between the population outflows and the per capita COVID-19 search frequency for each Chinese city in the study*. We hypothesized that well-informed individuals might be educated to follow the “stay-at-home” policy in a stricter manner. We correlated the outflow recovery rate with the per capita COVID-19-related search frequency in every city, and we evidenced the significance of the negative correlation, such that *R*^***^ = −30.96% (*N* = 307 and *p*-value= 3.01 × 10^−8^ < 0.0001).

**Figure 5.**
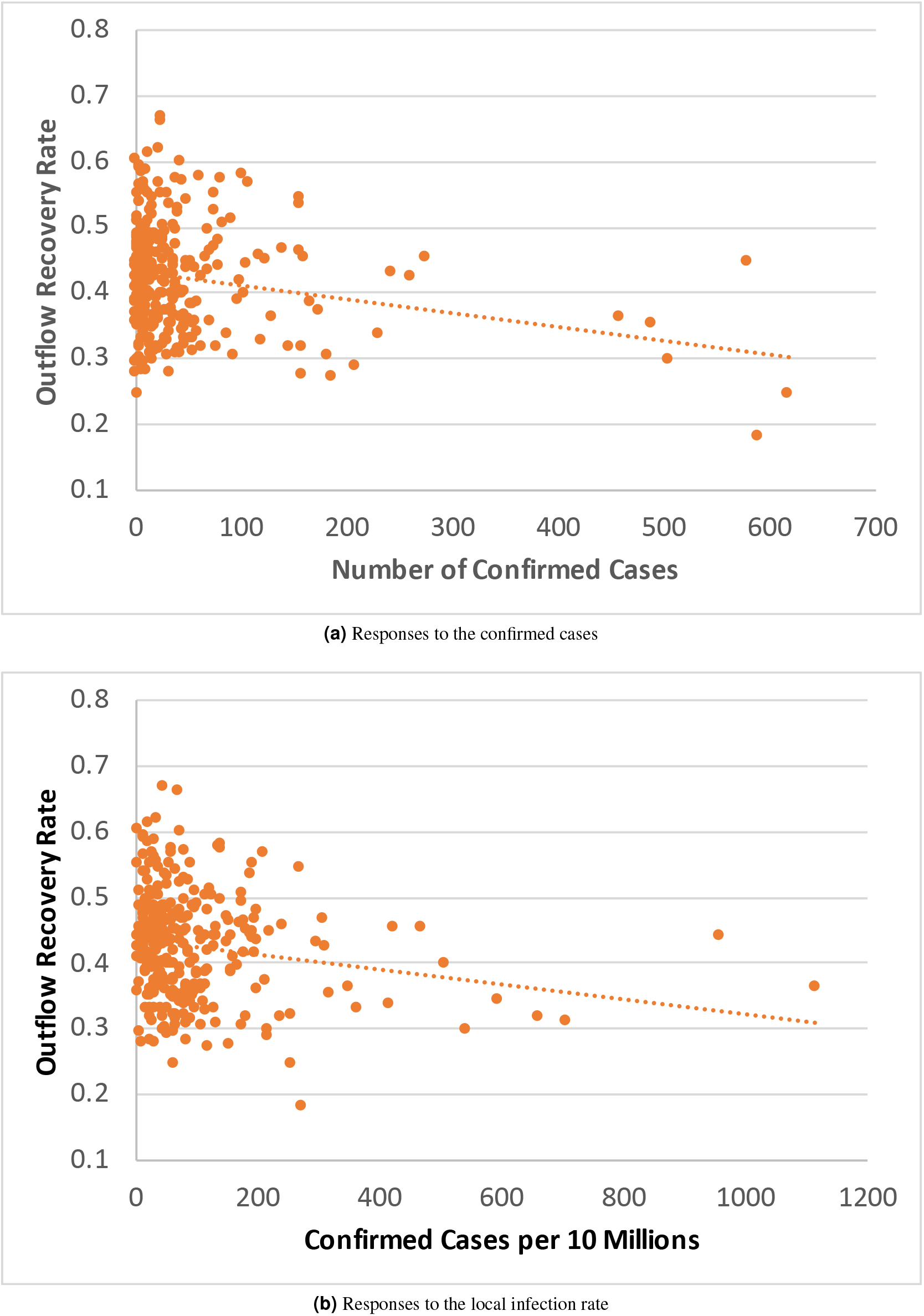
Understanding the outbound travels as the collective responses of populations to the infectious situation in every city. (Significant negative correlations have been found between outflows and infection situations.)

As both per capita COVID-19-related search frequency and outflows are correlated to the local infections, to avoid the possible threat to validity, we also carried out partial correlation analysis with the number of confirmed cases as the controlling variable. The result continues to be significant *R*^***^ = −26.23% (*N* = 307 and *p*-value = 3.16 × 10^−6^ < 0.0001). Similar partial correlation analysis has been done with local infection rate as the controlling variable, where we again evidenced the significance of correlation such that *R*^***^ = −26.47% (*N* = 307 and *p*-value= 2.57 × 10^−6^ < 0.0001). In addition to infections, yet another variable that would affect the significance is the overlaps of Baidu Maps and Baidu search user communities in different cities. Considering the coverage of Baidu users throughout the Chinese population, we follow the previous work^17,18^ to set this issue aside. Please see also Figure 6 for the visualization of the correlations.

**Figure 6.**
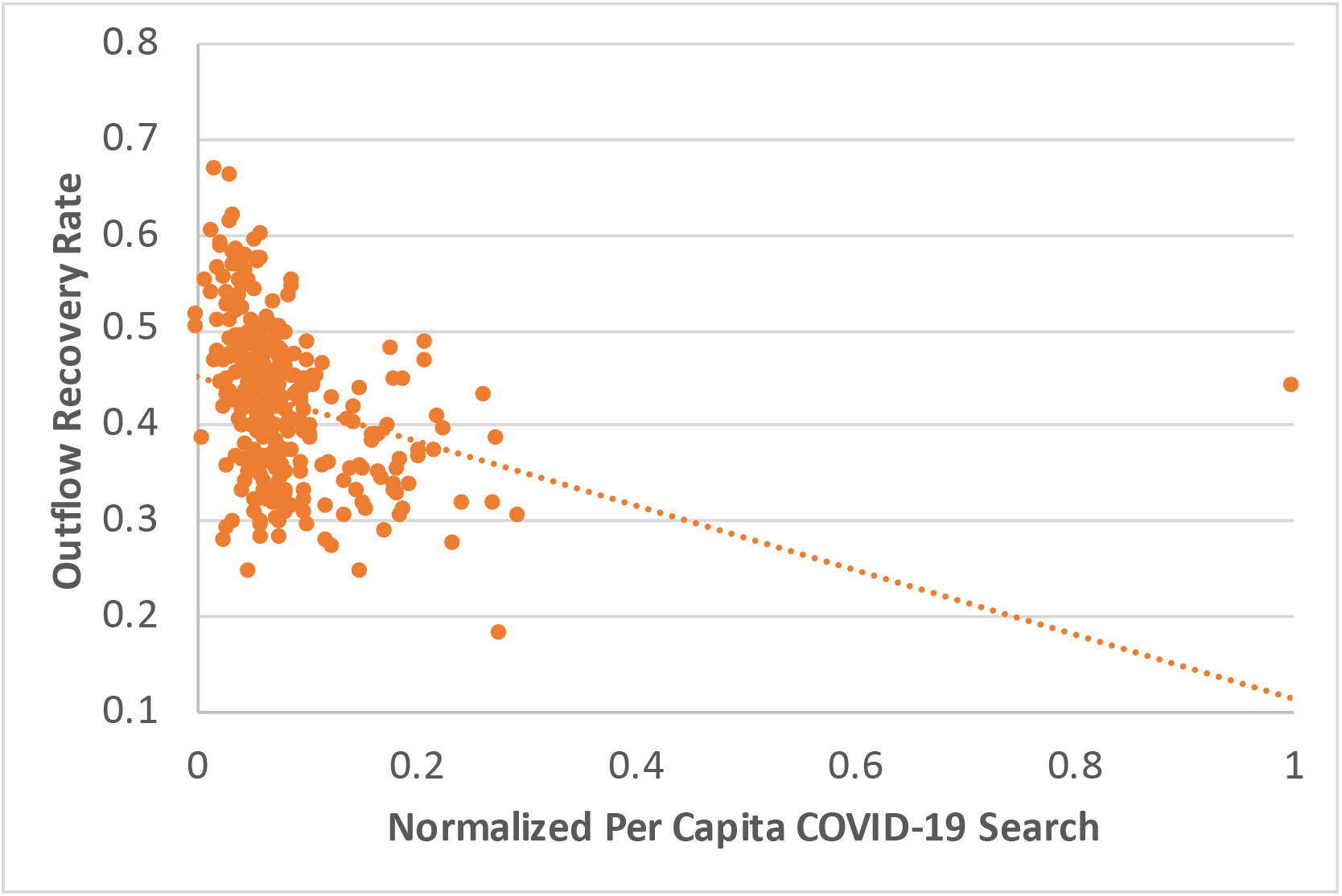
Understanding the interplay between the outbound travels and the COVID-19-related search in every city. (Significant negative correlations have been found between the per capita COVID-19-related search frequency of every city and the local outflow recovery rates.)

## Discussion and Conclusion

In this work, we re-examined the connection between inflows from Wuhan and the infections in every major Chinese city, using more concrete mobility data, and we tested three unique hypotheses, which provide us with novel understandings of the human responses to the COVID-19 pandemic, in both cyber and physical worlds. As the collective responses of the population to the emergencies, the demands of information from individuals are higher as the local infectious situation becomes worse. The more infectious cases in a city, the more the residents of the city would search online for pertinent information. However, people are not likely to escape from the cities where they live, even when the situations are critical. The worse the infectious situation in a city, the less likely residents are to travel beyond the city. Furthermore, well-informed individuals are more likely to travel less, even while the total travel intentions are still low compared to the prior year. The more the residents of a city have engaged searches pertaining to the pandemic, the more likely those residents are to refrain from travel. The implications of these correlations include that, the more that timely information acquisition is attainable to and attained by residents, the more stay-at-home orders are likely to be followed, even, or especially, when there are more cases in a city.

## Methods

To carry out the study, we collected four datasets of three categories as follows.

- **COVID-19-related search -** We extracted search queries that include 40 keywords and phrases related to COVID-19, such as novel coronavirus, SARS, respiratory diseases, vaccines, Shuang-Huang-Lian (a traditional Chinese medicine which was sold out all over the country and was believed symptom-relieving^19^), etc. We localized each query and categorized the queries by the location. Finally, for every major Chinese city (307 cities in total, with cities in Hubei Province excluded), we collected *(Dataset I)* the total number of COVID-19-related search queries during the period from 23 Jan 2020 to 31 Mar 2020. To protect the privacy of users, we did not track the user identities and kept them anonymous. Note that Baidu has been reported as the largest Chinese search engine in the world, with a 75% market share in Mainland China and 195 million average daily active users (around 13% of Chinese nationals) by the end of December 2019.^3^
- **Inter-city mobility -** We used anonymized localization request logs of Baidu Maps and detected inter-city mobility as the location change in two consequent localization requests. The transitions of inter-city mobility were categorized by the origin and destination (OD) pairs. In general, we collected *(Dataset II)* the inter-city transitions between major Chinese cities during the periods from 23 Jan 2020 (Wuhan lockdown) to 31 Mar 2020. As a reference, we also collected *(Dataset III)* the inter-city mobility traces in the same format for the same period around Chinese New Year of 2019 (from 3 Feb 2019 to 12 Apr 2019). Note that Baidu Maps is one of the largest Web mapping applications with over 340 million monthly active users worldwide by the end of December 2016.^4^
- **Open data on COVID-19 cases and consensus -** With respect to the previous work^7–11^, we also collected *(Dataset IV)* the number of confirmed infectious cases (up-to-date by 31 Mar 2020) for every major Chinese city from Baidu COVID-19 Page and the population of the city from Baidu Encyclopedia.

For the techniques and settings that have been used to perform data normalization, data visualization, and correlation analysis, please refer to the supplementary materials.

## Data Availability

All experiments in this paper were carried out using anonymous data and secure data analytics provided by Baidu Data Federation Platform (Baidu FedCube). For data accesses and usages, contact us via {fedcube, shubang}@baidu.com.

## Acknowledgement

All experiments in this paper were carried out using anonymous data and secure data analytics provided by Baidu Data Federation Platform (Baidu FedCube). For data accesses and usages, contact us through http://shubang.baidu.com/page/page_en.html.

## Author contributions statement

H.X. formulated the research problems and drafted the manuscript. H.X., J.L., and J.H. collected data from Baidu, conducted the experiments, and performed data analysis. S.H., H.A., and Q.K. collected the consensus data and carried out the data visualization. Y.L. managed the user privacy protocol and was in charge of data anonymization. D.D. and H.W. proposed the research, coordinated the research efforts, and oversaw the whole research process.

## A Supplementary Materials

Here we introduce the details on data normalization, data visualization, and procedures used for correlation analysis.

### A.1 Data Normalization

To visualize the data while preserving privacy on Baidu users, we plotted figures using normalized data, as follows.

- *Normalized volumes of population inflows from Wuhan* and *normalized volumes of COVID-19-related search -* We normalized the plots in Figures 1, 3 and 8 to the proportional values (ranging from 0 to 1) using the maximum values appeared in the plots as 1.0.
- *Normalized Inflows from Wuhan -* In Figure 2, we visualized the correlation between the number of confirmed infection cases and the normalized inflows from Wuhan. Given the volumes of population inflows from Wuhan (1 Jan to 23 Jan 2020) for every city in the study, we normalize the data using *min-max scaling*. Such that, given the overall volumes of inflows (1 Jan to 23 Jan 2020) from Wuhan to each of the 307 cities in the study, we first found the maximum and minimum of the inflow volumes as Inflow_max_ and Inflow_min_ respectively, then we normalize inflow volume of every city as

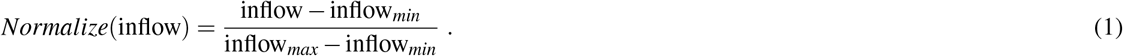 In this way, the volumes of population inflows from Wuhan to every city in the study have been curved into the range from 0 to 1 proportionally.
- *Normalized Per Capita Search (Log) –* In Figure 7, Figure 4, and Figure 6, we visualized the correlations of the per capita COVID-19-related search frequency of every city in the study to the local population size, infection situations, and volumes of population outflows respectively. For better visualization, we used the *min-max scaling* over *10-Base logarithm* of the per capita COVID-19-related search. Such that, given the per capita COVID-19-related search frequency of every city, we first find the maximum and minmum values as search*_max_* and search*_min_* respectively, then we normalize them as

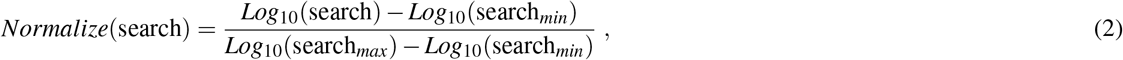

where *Log*_10_ (·) refers to a 10-base logarithmic function. In this way, the logarithms of per capita COVID-19-related search frequency of every city in the study have been curved into the range from 0 to 1 proportionally.
- *Outflow Recovery Rates -* We have already defined the calculation of *outflow recovery rates* in the paragraphs of Result III. More specifically, for every city in the study, given the volumes of population outflows from the city in the comparable periods of 2019 and 2020 as outflow_2019_ and outflow_2020_ respectively, we calculate the *outflow recovery rate* of the city as outflow_2020_/outflow_2019_. In this way, we could project the volumes of outflow in 2020 back to the comparable period of 2019, and estimate to what degree each city’s traffic has been recovered.

### A.2 Correlation Analysis

We conducted two types of correlation analysis in our study. To understand the correlation between two random variables (bivariate correlation analysis), we estimated the Pearson correlation coefficients and performed the Student’s T-test (two tails) for the significance test. We considered four levels of significance (< 0.0001, ≥ 0.0001 but < 0.001, ≥ 0.001 but < 0.01, and ≥ 0.01 but < 0.05) for statistical hypothesis testing.

Furthermore, we sometimes needed to test the degree of association between two random variables, with the effect of a set of random controlling variables removed. For example, in Result IV, we tested the significance of the correlation between search behaviors and mobility, while removing the effects of local infections as the controlling variable. We thus performed the partial correlation analysis. More specifically, given three random variables *X*, *Y*, and *Z*, we first estimated the Pearson correlation coefficients between any two of these three variables as Corr_X Y_, Corr_YZ_, and Corr_X Z_ respectively. We then estimated the partial correlation coefficient that measures the correlation between *X* and *Y* with the effect of *Z* removed as

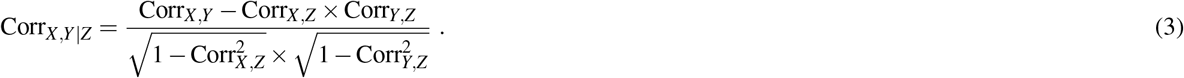

We follow the standard procedure of Student’s T-test for partial correlation coefficients to test the significance of Corr_X_ *_Y_*_|Z_ with *p*-value.

In general, we performed the correlation analysis using the formal settings of significance tests. We understand there might exist better methods to analyze the data and interpret the correlations.

**Figure 7.**
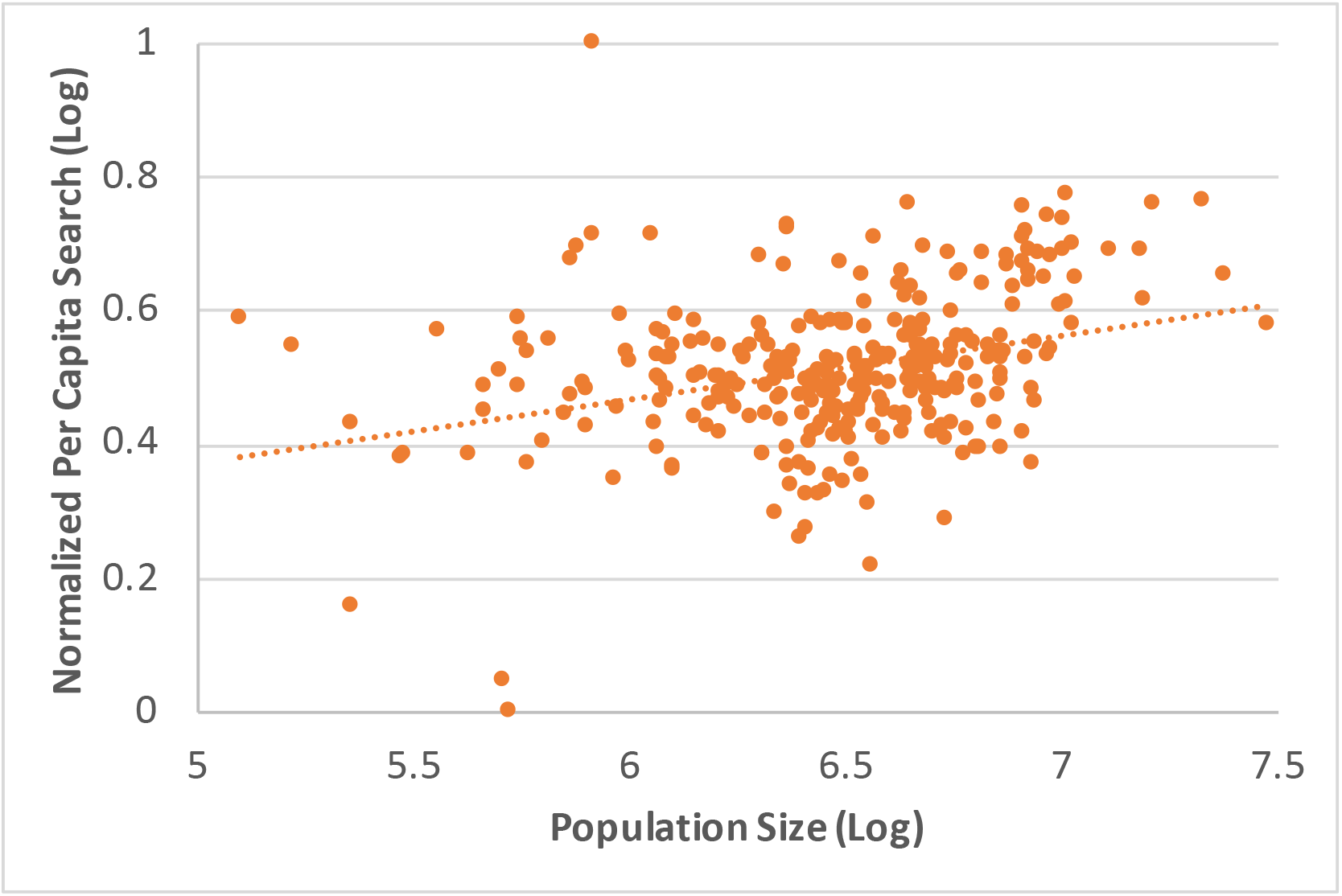
As a response to the emergencies, individuals are likely to search for COVID-19-related information more frequently in relatively larger cities with greater populations. (We thus need to include the population size of every city as a controlling factor to analyze the search data.)

### A.3 Correlations have been evidenced between the per capita COVID-19-related search activities and the population size of the city in major Chinese cities

We consider the size of population in every city as an important factor that might affect the results of correlation analysis. We thus correlated the per capita COVID-19-related search frequency with the population size of every major city in Mainland China. Figure 7 illustrated the log-log plots of the correlation between the two factors.

### A.4 Population Inflows and Outflows between Wuhan and Beijing, Shanghai, Guangzhou, and Shenzhen in 2019

The 2019 peak of population inflows from Wuhan to the four largest cities in Mainland China was in 7 Apr 2019 (not the Chinese New Year). To validate our observations, we checked out the dates and found that 5 Apr 2019 is the Qingming Festival and there were public holidays from 5 Apr 2019 to 7 Apr 2019. Qingming Festival is the Tomb-Sweeping Day in Chinese traditions. It is reasonable to consider that the Beijing, Shanghai, Guangzhou, and Shenzhen residents whose hometown is Wuhan (or whose hometown is in surrounding areas) would travel back to Wuhan for the festival and related activities. Figures 8(a) and (b) demonstrate the comparison between the volumes of population inflows from Wuhan and outflows to Wuhan respectively. From 3 Apr 2019 to 5 Apr 2019, we observed large volumes of population outflows from the four cities to Wuhan. Then the round trips back to the four cities (inflows from Wuhan to the four cities) happened immediately in the following days, since the length of public holidays was short. Thus it makes sense that the 2019 peak of population inflows from Wuhan to the four cities arrived on 7 Apr 2019.

**Figure 8.**
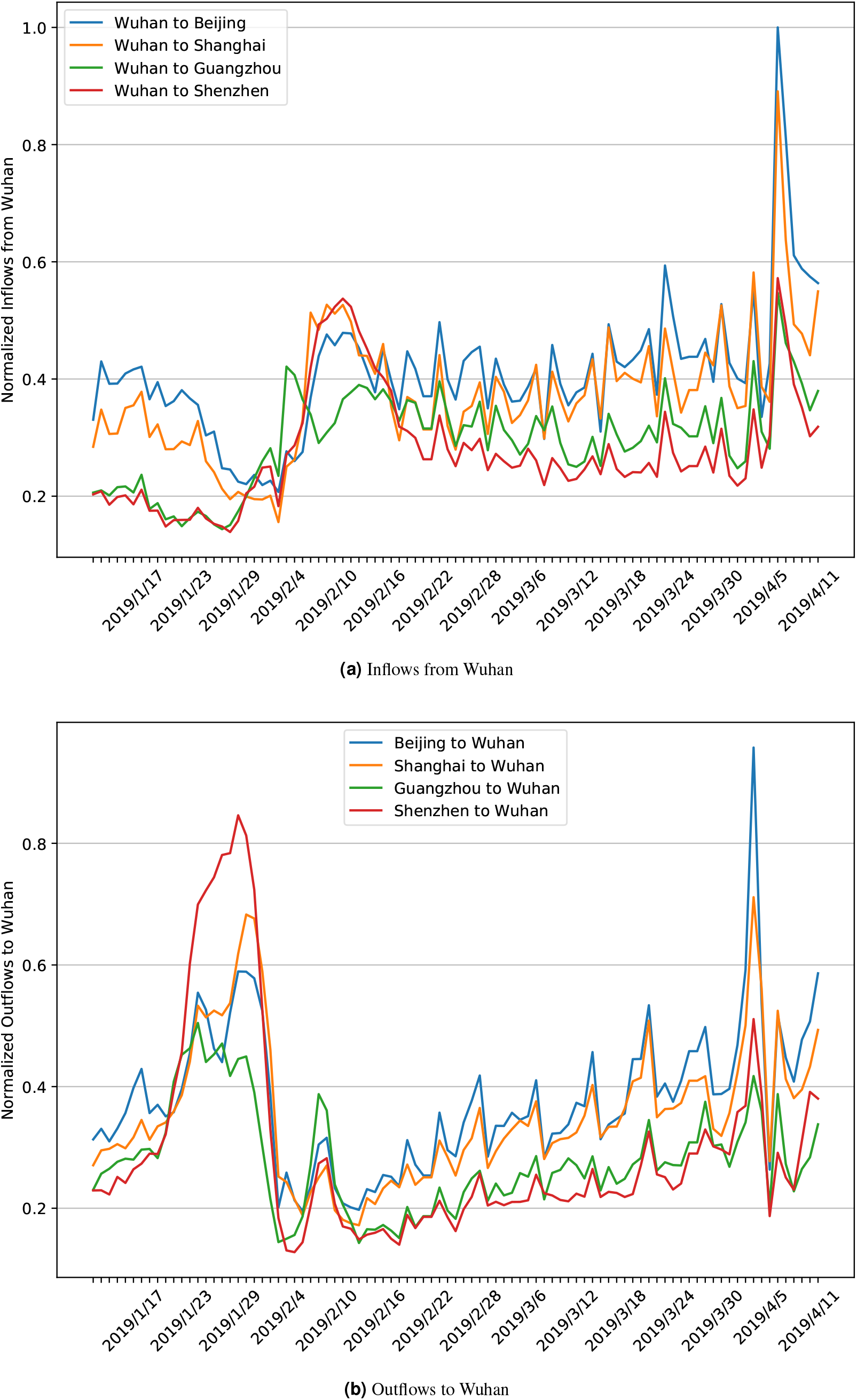
A comparison between the population inflows from Wuhan and the population outflows to Wuhan

### A.5 The spatial distribution of COVID-19-related searches over major cities of Mainland China

Figure 9 presents the distribution of COVID-19-related searches evolving over major cities in Mainland China. We illustrated the spatial distributions of the normalized volumes of COVID-19-related searches over major Chinese cities in four key dates—15 January 2020 (early stage of infections), 23 January 2020 (Wuhan lockdown), 31 January 2020 (the peak of nationwide search volumes), and 31 March 2020 (cool down of the searches).

**Figure 9.**
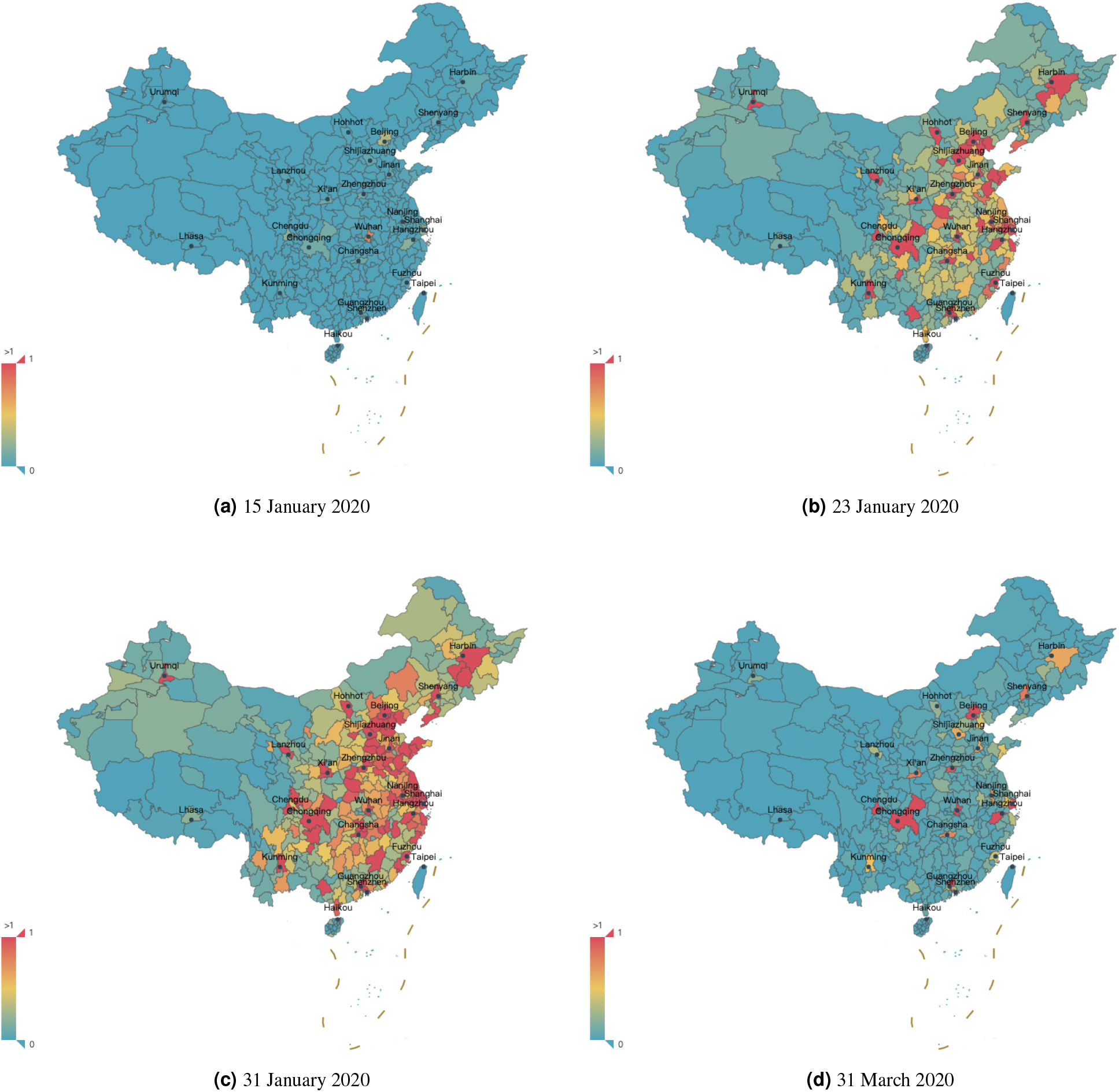
The spatial distribution of COVID-19-related searches (normalized volumes) over major cities of Mainland China in the study.

### Footnotes

1 Baidu Migration - http://qianxi.baidu.com/

2 Baidu COVID-19 Page - https://voice.baidu.com/act/newpneumonia/newpneumonia/

3 http://ir.baidu.com/static-files/9e8f63e2-f22b-4cba-8e65-9120ba40fa35

4 http://ir.baidu.com/static-files/e249a0f8-082a-4f8a-b60d-7417fa2f8e7e

